# Ambient Heat and Age-dependent Blood Pressure Responses in 184,000 U.S. Adults

**DOI:** 10.64898/2026.11.25.26369026

**Authors:** Shuai Yang, David A. Sbarra, Yann C. Klimentidis, Heidi E. Brown

## Abstract

**Aims:** Extreme heat challenges cardiovascular regulation, particularly in older adults. Although blood pressure is generally lower in warmer weather, short-term within-person associations across the temperature distribution remain incompletely characterised.

**Methods:** We linked repeated outpatient blood-pressure measurements from the All of Us Research Program to hourly ERA5-Land weather data aggregated to local calendar days. The primary exposure was maximum temperature during the complete local day preceding each visit. Models with participant and exact calendar-date fixed effects included 2,279,955 person-days from 181,865 participants for systolic blood pressure and 2,266,080 person-days from 181,770 participants for diastolic blood pressure.

**Results:** Compared with a preceding-day maximum temperature of 18.4 °C (50th percentile), a temperature of 31.3 °C (95th percentile) was associated with lower systolic blood pressure (−0.66 mmHg; 95% confidence interval [CI], −0.77 to −0.55) and diastolic blood pressure (−0.28 mmHg; 95% CI, −0.35 to −0.21). Associations extended through the 99th percentile. The systolic difference between these two temperatures became 0.20 mmHg more negative per decade of age and reached −0.99 mmHg at age 75. Social-material vulnerability and long-term climate showed little evidence of modifying the associations, and the inverse associations persisted after adjustment for wildfire smoke. In secondary analyses, visit-to-visit blood-pressure variability showed little overall association with heat, although the heat-associated difference in absolute changes between successive systolic readings increased by 0.05 mmHg per decade of age.

**Conclusion:** Preceding-day heat was associated with modestly lower outpatient blood pressure within individuals, with stronger associations at older ages.

**Lay summary:** We studied how outdoor heat relates to blood pressure measured at medical clinic visits and found slightly lower readings after hotter days, particularly among older adults.

- Following a very warm day (31.3 °C) rather than a milder day (18.4 °C), average systolic blood pressure was 0.66 mmHg lower and diastolic blood pressure was 0.28 mmHg lower.
- The difference in systolic blood pressure increased with age, reaching approximately 1 mmHg at age 75; these small associations do not mean that heat is protective for cardiovascular health.

Graphical abstract. Model-estimated differences in systolic blood pressure (SBP) and diastolic blood pressure (DBP) comparing preceding-day maximum temperatures of 31.3 °C and 18.4 °C, overall and at ages 35, 55 and 75 years. Age-specific estimates are from the continuous-age models. Error bars indicate 95% confidence intervals. Alt text: A summary of the All of Us outpatient study showing progressively larger inverse temperature–blood-pressure associations at older ages.

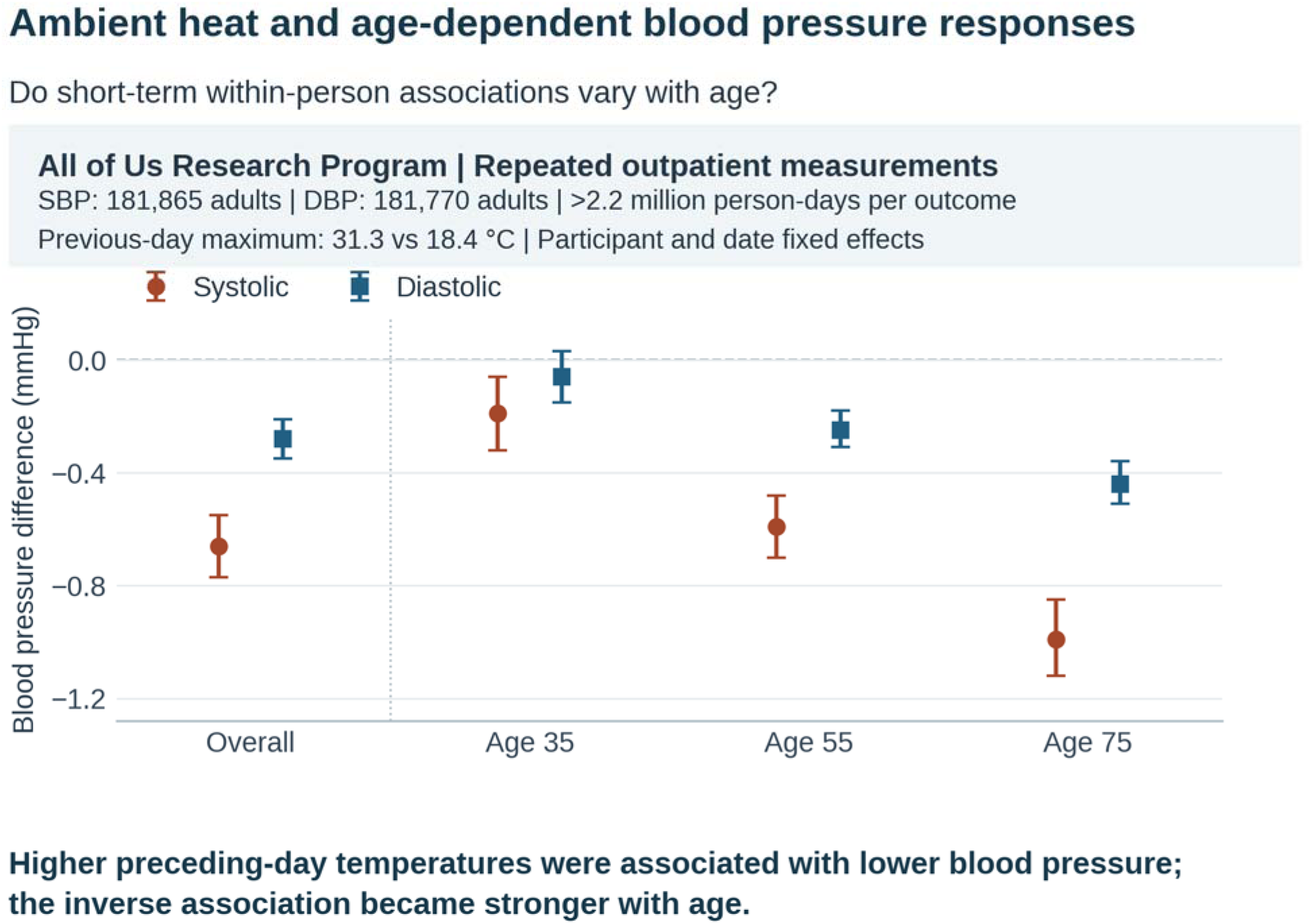

## Introduction

Extreme heat is an increasingly important cardiovascular hazard. Short-term exposure is associated with cardiovascular morbidity and mortality, particularly in older adults and people with cardiovascular disease (Liu et al., 2022). Heat dissipation increases skin blood flow and promotes sweating. Peripheral vasodilation reduces vascular resistance, while fluid loss through sweating can lower circulating volume and alter cardiac loading, making blood pressure informative about cardiovascular responses to heat (Chaseling et al., 2021; Cottle et al., 2025; Crandall & González-Alonso, 2010).

Laboratory studies have documented changes in stroke volume and cardiac filling during heat exposure (Foster et al., 2025; Meade et al., 2025). Epidemiological studies also show that blood pressure tends to be higher in colder weather and lower in warmer weather (Wang et al., 2017; Gronlund et al., 2018; Kent et al., 2011). Seasonal comparisons, however, cannot establish how a person’s blood pressure differs following short-term changes in local temperature.

In more than 4.2 million US community-health-centre encounters, McGrath et al. (2026) reported approximately 0.7 mmHg lower systolic and 0.3 mmHg lower diastolic blood pressure during same-day extreme heat. The systolic difference was 1.1 mmHg among adults aged 65 years or older. Their study defined heat using location-specific thresholds and reported the strongest lagged associations on the day of exposure and the following day. These observations provide an independent outpatient benchmark and support examining recent heat exposure. A continuous temperature-response curve can further show whether the inverse association persists into the hottest conditions; other repeated-measures research has reported higher systolic pressure during heatwave warnings among older adults with hypertension (Zhang et al., 2026). McGrath et al. (2026) used patient- and facility-level random intercepts and identified a fully within-patient fixed-effects design as a useful next step.

Age may shape this relationship. Older adults can face greater circulatory demands during heat exposure as cardiovascular and thermoregulatory reserve declines (Cottle et al., 2025). Controlled experiments have documented age-related differences in cardiac responses to heat (Foster et al., 2025). Treatments affecting vascular tone or fluid balance may provide additional context for these responses. Blood pressure also varies between clinical encounters, and visit-to-visit variability has been associated with cardiovascular outcomes beyond mean blood pressure (Lukitasari et al., 2026).

The relationship between outdoor temperature and blood pressure may also vary with living conditions and local climate. Housing and access to cooling can influence personal heat exposure, while long-term climate may shape adaptation to hot weather (Chu et al., 2026). In addition, hot weather can coincide with wildfire smoke, which has been linked to cardiovascular stress and concurrent heat-related health burdens (Heaney et al., 2022; Jones-Ngo et al., 2025; Walker et al., 2025). These contexts motivate examining the consistency of the acute temperature association across social environments and during smoke exposure.

Using longitudinal electronic health records (EHRs), surveys and generalised residential geography from the All of Us Research Program (All of Us Research Program Investigators, 2019), we examined the outpatient temperature–blood-pressure association in a separate cohort. Participant fixed effects accounted for measured and unmeasured characteristics that remain stable within a person, and exact calendar-date fixed effects accounted for influences shared nationally on a given date. We characterised the continuous temperature-response curve across adulthood and examined visit-to-visit variability as a secondary outcome. We also assessed modification by social-material vulnerability and long-term local climate, and examined whether the association persisted after accounting for wildfire smoke and whether the two exposures interacted. The complete local day before each visit provided an exposure window that ended before the measurement date.

## Methods

### Study design and data source

We conducted a national repeated-measures observational analysis using the All of Us Research Program Controlled Tier Curated Data Repository version 9 (CDR v9; C2025Q4R6), with a cutoff date of January 1, 2025. The blood-pressure study window was November 1, 2021 through December 31, 2024. Analyses were conducted in the secure All of Us Researcher Workbench. Controlled Tier event dates permit linkage of electronic health record measurements with daily environmental conditions (All of Us Research Program Investigators, 2019).

### Study population and analytic samples

Adults contributed to the primary analysis if they were at least 18 years old on the measurement date, had a generalised residential three-digit ZIP-code prefix and had at least two eligible blood-pressure measurement days for the modelled outcome with required weather covariates. The primary systolic and diastolic models included 181,865 and 181,770 participants, respectively (Table 1). Analyses of social-material vulnerability additionally required a scoreable survey index date and were limited to subsequent blood-pressure measurements. Smoke analyses were restricted to dates with verified coverage of the Stanford Environmental Change and Human Outcomes (ECHO) data product. These restrictions defined different analytic panels, with outcome-specific counts in Supplementary Table S1.

**Table 1.**
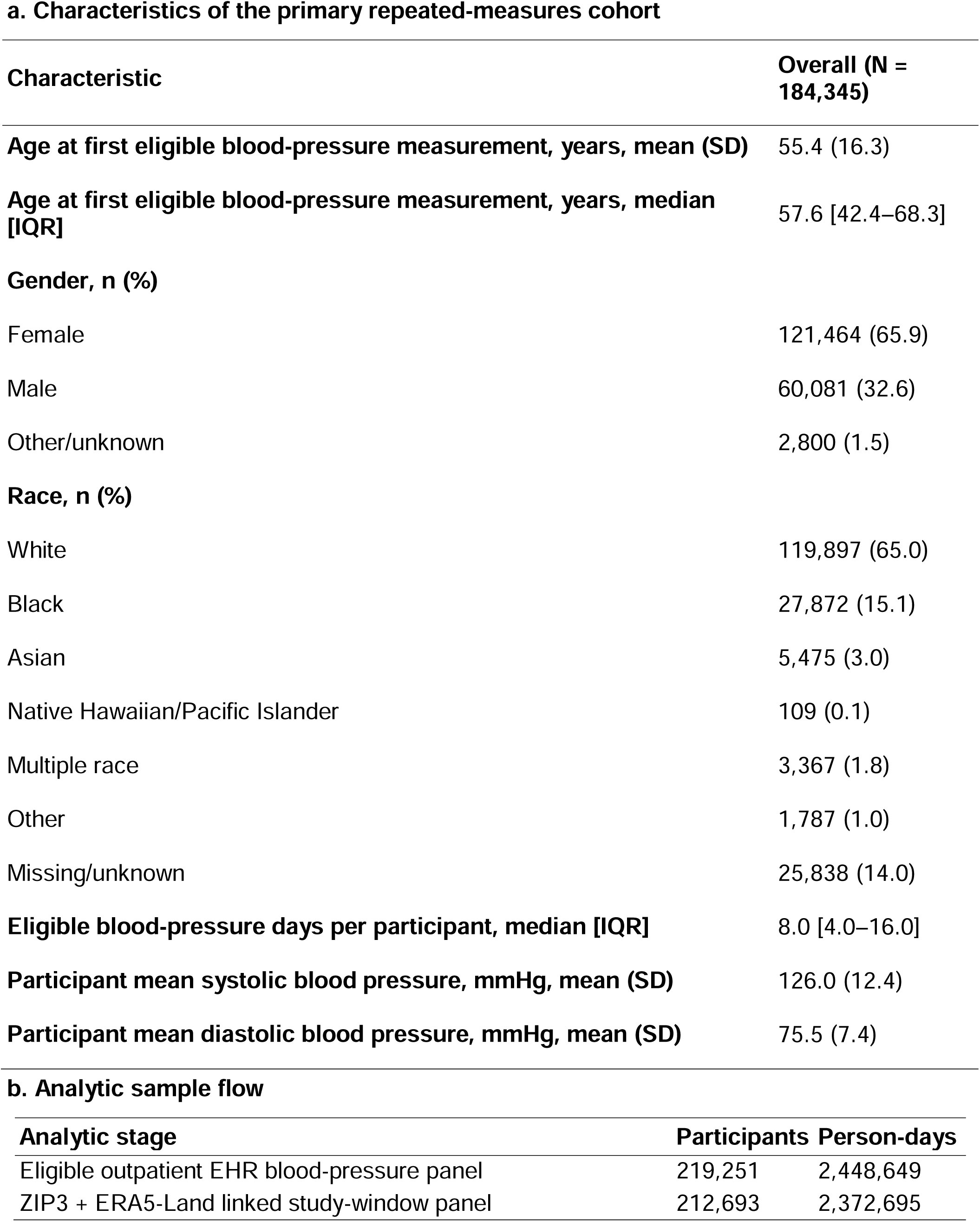
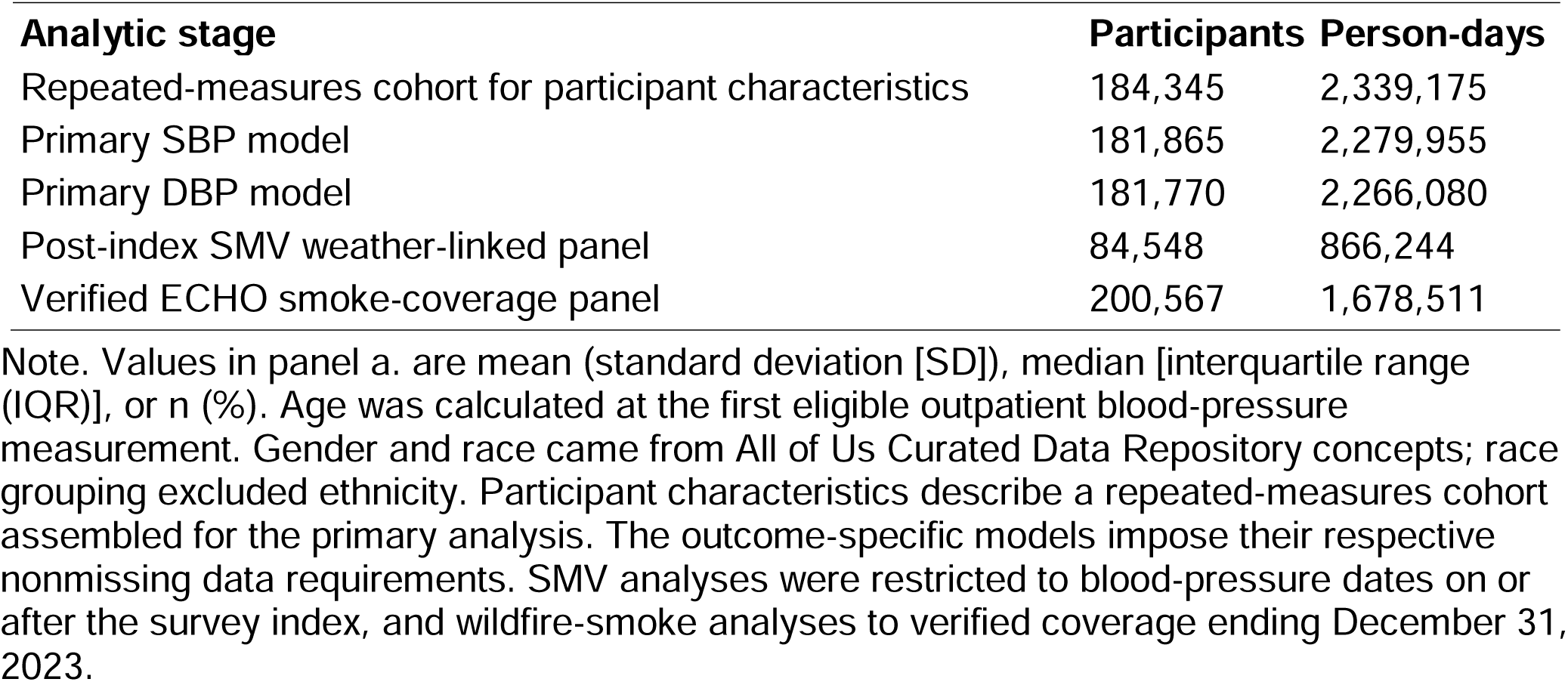
Participant characteristics and analytic sample flow.

| **a. Characteristics of the primary repeated-measures cohort** |  |  |
| **Characteristic** | **Overall (N = 184,345)** |  |
|---|---|---|
| **Age at first eligible blood-pressure measurement, years, mean (SD)** | 55.4 (16.3) |  |
| **Age at first eligible blood-pressure measurement, years, median [IQR]** | 57.6 [42.4–68.3] |  |
| **Gender, n (%)** |  |  |
| Female | 121,464 (65.9) |  |
| Male | 60,081 (32.6) |  |
| Other/unknown | 2,800 (1.5) |  |
| **Race, n (%)** |  |  |
| White | 119,897 (65.0) |  |
| Black | 27,872 (15.1) |  |
| Asian | 5,475 (3.0) |  |
| Native Hawaiian/Pacific Islander | 109 (0.1) |  |
| Multiple race | 3,367 (1.8) |  |
| Other | 1,787 (1.0) |  |
| Missing/unknown | 25,838 (14.0) |  |
| **Eligible blood-pressure days per participant, median [IQR]** | 8.0 [4.0–16.0] |  |
| **Participant mean systolic blood pressure, mmHg, mean (SD)** | 126.0 (12.4) |  |
| **Participant mean diastolic blood pressure, mmHg, mean (SD)** | 75.5 (7.4) |  |
| **b. Analytic sample flow** |  |  |
| **Analytic stage** | **Participants** | **Person-days** |
| Eligible outpatient EHR blood-pressure panel | 219,251 | 2,448,649 |
| ZIP3 + ERA5-Land linked study-window panel | 212,693 | 2,372,695 |
| **Analytic stage** | **Participants** | **Person-days** |
| Repeated-measures cohort for participant characteristics | 184,345 | 2,339,175 |
| Primary SBP model | 181,865 | 2,279,955 |
| Primary DBP model | 181,770 | 2,266,080 |
| Post-index SMV weather-linked panel | 84,548 | 866,244 |
| Verified ECHO smoke-coverage panel | 200,567 | 1,678,511 |

Note. Values in panel a. are mean (standard deviation [SD]), median [interquartile range (IQR)], or n (%). Age was calculated at the first eligible outpatient blood-pressure measurement. Gender and race came from All of Us Curated Data Repository concepts; race grouping excluded ethnicity. Participant characteristics describe a repeated-measures cohort assembled for the primary analysis. The outcome-specific models impose their respective nonmissing data requirements. SMV analyses were restricted to blood-pressure dates on or after the survey index, and wildfire-smoke analyses to verified coverage ending December 31, 2023.

### Residential geography and environmental linkage

Environmental exposures were assigned with the generalised three-digit ZIP-code prefix (ZIP3) available in the Controlled Tier. We used the most recent generalised residential ZIP3 record in CDR v9 and retained the date of that record. A sensitivity analysis restricted clinical measurements to within 365 days of the residential record. Public environmental data were processed outside the Researcher Workbench and imported only after aggregation to ZIP3-day level.

### Meteorologic data

Acute meteorology came from hourly ERA5-Land reanalysis data (Muñoz-Sabater et al., 2021). A fixed ZIP3-to-grid mapping linked each ZIP3 to an ERA5-Land grid cell and an Internet Assigned Numbers Authority time zone. Hourly Coordinated Universal Time timestamps were converted to that local time zone before daily aggregation. Daily maximum temperature (T_max_), mean temperature and minimum temperature were calculated from hourly 2-m air temperature. Mean 2-m dew-point temperature and total precipitation were derived over the same local day. The algorithm explicitly handled 23-hour and 25-hour daylight-saving days. The resulting ZIP3-day dataset contained 1,046,628 ZIP3-day rows, representing 881 ZIP3s from October 1, 2021 through December 31, 2024. Processing details and quality checks are provided in the Supplementary Methods and Supplementary Table S1.

### Repeated blood-pressure outcomes

We extracted systolic blood pressure (SBP) and diastolic blood pressure (DBP) from standard Observational Medical Outcomes Partnership measurement concepts. Measurements taken as part of All of Us enrolment physical assessments were excluded. The primary phenotype used outpatient and ambulatory encounters; readings outside 70–250 mmHg for SBP and 40–150 mmHg for DBP were removed. When a participant had more than one eligible reading of a given type on a day, we averaged those values. Pulse pressure and mean arterial pressure were secondary outcomes for days with paired valid measurements in which systolic pressure exceeded diastolic pressure.

### Visit-to-visit blood-pressure variability

For the secondary visit-to-visit variability outcome, we calculated the absolute successive difference (ASD), the absolute change between consecutive eligible blood-pressure measurements of the same type. For example, systolic readings of 130 and 120 mmHg give an ASD of 10 mmHg. This interval-specific contribution to average real variability can change within a participant, making it suitable for participant fixed-effects models. We retained intervals of up to 365 days, required at least two eligible intervals per participant and adjusted for the natural logarithm of one plus the number of days between readings. A maximum 90-day interval was examined in sensitivity analysis. Participant-level standard deviation, coefficient of variation and average real variability were calculated descriptively; details are in the Supplementary Methods.

### Primary temperature exposure

Maximum temperature (T_max_) during the complete local calendar day immediately preceding the blood-pressure measurement was the primary exposure (local day t−1). The measurement-date temperature maximum could occur after the clinic visit, so we reserved same-day exposure for sensitivity analysis. We modelled the preceding-day T_max_ using a four-degree-of-freedom natural cubic spline; knots and boundary knots were derived once from the primary exposure distribution and reused across models. The median T_max_ was 18.45 °C, the 95th percentile was 31.28 °C, and the 99th percentile was 34.92 °C.

### Temperature contrasts and weather covariates

We summarised the continuous fitted curve using a primary contrast between the 95th (P95) and 50th (P50; median) temperature percentiles. The 5th percentile (P5) versus P50 was a cold-side benchmark, and the 99th percentile (P99) versus P50 described the extreme upper tail. These contrasts represent model-estimated differences at selected temperatures along the fitted curve. Previous-local-day mean dew-point temperature and precipitation were time-varying covariates. Mean T_max_ over local days t−1 and t−2 was a secondary exposure window.

### Long-term climate context

Long-term climate context was represented by a ZIP3-specific cooling-degree-day (CDD) normal derived from 1991-2020 daily mean-temperature normals from the Parameter-elevation Regressions on Independent Slopes Model (PRISM), using a 65 °F (18.3 °C) base (Daly et al., 2008). Higher values indicate historically warmer climates. The acute ERA5-Land exposure and the long-term PRISM climate measure were kept conceptually separate.

### Social-material vulnerability

We calculated a continuous social-material vulnerability (SMV) index using established All of Us survey measures of social circumstances alongside an area-level deprivation index supplied in the CDR v9 ZIP3 socioeconomic status table. The index is derived from American Community Survey indicators of material deprivation using the approach of Brokamp et al. (2019); the source is documented by the All of Us Research Program (2026). The components assessed loneliness, functional rurality or access to resources, social support, neighbourhood conditions, material hardship and socioeconomic resources. We standardised available components, coded higher scores as greater vulnerability and calculated their equal-weight mean when at least three were observed; the resulting score was standardised again. The composite was constructed for this study using existing survey domains. The survey index date and individual component definitions are provided in the Supplementary Methods.

### Wildfire-smoke exposure

Wildfire-specific fine particulate matter with aerodynamic diameter 2.5 µm or smaller (PM_2.5_) came from the Stanford Environmental Change and Human Outcomes daily smoke product based on the Childs et al. framework (Childs et al., 2022). This product provides modelled daily wildfire-attributable ground-level PM_2.5_ estimates for ZIP Code Tabulation Areas. ZIP Code Tabulation Area (ZCTA)-day estimates were aggregated to ZIP3 using 2022 American Community Survey population weights. Within verified coverage, locations without a wildfire-smoke record contributed zero wildfire-attributable PM_2.5_. Dates outside verified coverage remained missing. The available product extended through December 31, 2023.

## Statistical analysis

For our first research question, we estimated the association between previous-day T_max_ and systolic or diastolic blood pressure using two-way fixed-effects linear models with participant and exact calendar-date effects (Gunasekara et al., 2014). Participant effects accounted for stable individual characteristics, and calendar-date effects captured conditions shared nationally on a given date. The continuous four-degree-of-freedom temperature spline was included with previous-local-day dew point and precipitation. Standard errors were clustered by participant. Model-estimated differences at P95 versus P50 were the primary contrasts; P5 and P99 served as benchmarks. Holm correction was applied across the two primary outcome tests.

To assess age modification, we fitted two separate models. The categorical model included indicators for age at each measurement (<45, 45–64 or ≥65 years), their main effects and interactions with the temperature spline. We retained the group main effects because participants could cross an age boundary during follow-up. The continuous-age model instead interacted the common spline with age at each measurement and estimated temperature contrasts at ages 35, 55 and 75 years, and changes per decade. Joint Wald tests assessed the respective interaction terms. Gender- and race-specific analyses were secondary. Definitions and temperature-support checks appear in Supplementary Table S2.

For the second research question, SMV entered the model as a continuous score interacting with each temperature-spline term. To make the modification interpretable, we reported the difference between the model-estimated P95-versus-P50 contrasts at the 90th and 10th percentiles of SMV, with joint Wald tests for the full interaction. Temperature reference points came from the post-survey-index sample; common support was checked in the lowest and highest SMV quintiles (Supplementary Table S3).

We evaluated whether the acute temperature association differed across historically warmer and cooler ZIP3 areas. Each participant was assigned a ZIP3-specific 1991–2020 cooling-degree-day normal, which we standardised and interacted with the temperature spline. We compared fitted curves across the 10^th^, 50^th^ and 90^th^ percentiles of the climate distribution and used joint Wald tests to evaluate heterogeneity (Supplementary Table S4).

Finally, to assess interaction with smoke, we estimated smoke associations with blood pressure while adjusting for a three-degree-of-freedom spline of same-day mean temperature, dew point and precipitation. A mutually adjusted model combined wildfire-specific fine particulate matter with the primary previous-day T_max_ spline. We tested whether the temperature curve differed with smoke exposure through spline-by-smoke interactions. Separate same-local-day compound categories compared heat-only, smoke-only and concurrent exposure with neither exposure (Supplementary Table S5).

We assessed robustness of the mean-blood-pressure results to spline flexibility, heat exposure timing and study-panel definitions. Temperature windows averaged the preceding two or three complete local days; alternative specifications used same-day T_max_, previous-day mean or minimum temperature, and three- or five-degree-of-freedom splines. We also evaluated stronger repeated-measure requirements, equal-person weights, warm-season restriction, recent residential ZIP3 information, ZIP3-by-year-month fixed effects and multiway clustered standard errors. The SMV analysis was additionally repeated using all otherwise eligible blood-pressure dates, including those before its survey index date. Same-day T_max_ remained a temporally non-strict sensitivity analysis.

For the secondary variability question, we applied the same preceding-day temperature spline to ASD and tested its interaction with continuous age. The SMV modification analysis constructed ASD only from post-index blood-pressure measurements, ensuring that both readings in each interval followed the survey date. We applied Holm correction separately to the two overall variability tests and the two per-decade age-modification tests. Full diagnostic and sensitivity specifications are provided in the Supplementary Methods and tables.

Analyses were performed in R 4.5.3 within the All of Us Workbench using the *fixest* package (Bergé et al., 2026). We checked the ERA5-Land ZIP3-day data for duplicate ZIP3-date records, complete local-day coverage and meteorologic plausibility and verified spline construction before model fitting. No additional missing-data imputation was performed. Additional processing details are provided in the Supplementary Methods.

## Results

### Cohort and exposure distribution

The outpatient EHR extraction contained 2,448,649 blood-pressure person-days from 219,251 adults. Linking residential geography to ERA5-Land weather yielded 2,372,695 person-days from 212,693 people. Participant characteristics are available for a repeated-measures cohort of 184,345 adults contributing 2,339,175 eligible days (Table 1). Their mean age at first eligible measurement was 55.4 years (standard deviation, 16.3); 65.9% were female, and the median number of eligible blood-pressure days during the November 2021–December 2024 study period was 8 (interquartile range, 4–16). The primary systolic model included 2,279,955 person-days from 181,865 participants; the primary diastolic model included 2,266,080 person-days from 181,770 participants. For the analysis with SMV, the post-index weather-linked panel included 866,244 person-days from 84,548 people. The smoke-coverage panel included 1,678,511 person-days from 200,567 participants. Outcome-specific sample sizes are detailed in Supplementary Table S1.

### Research question 1: Short-term temperature and blood-pressure responses

We fitted previous-day maximum temperature as a continuous spline and report selected contrasts along that curve. At 31.28 °C (P95) compared with 18.45 °C (P50), systolic blood pressure was 0.66 mmHg lower (95% confidence interval [CI], −0.77 to −0.55; Holm-adjusted P<.001) and diastolic blood pressure was 0.28 mmHg lower (95% CI, −0.35 to −0.21; Holm-adjusted P<.001) (Table 2). The fitted curves declined above the median (Figure 1). P99-versus-P50 contrasts were −0.84 and −0.38 mmHg, respectively; overall P5-versus-P50 contrasts were −0.06 and 0.04 mmHg.

**Table 2.**
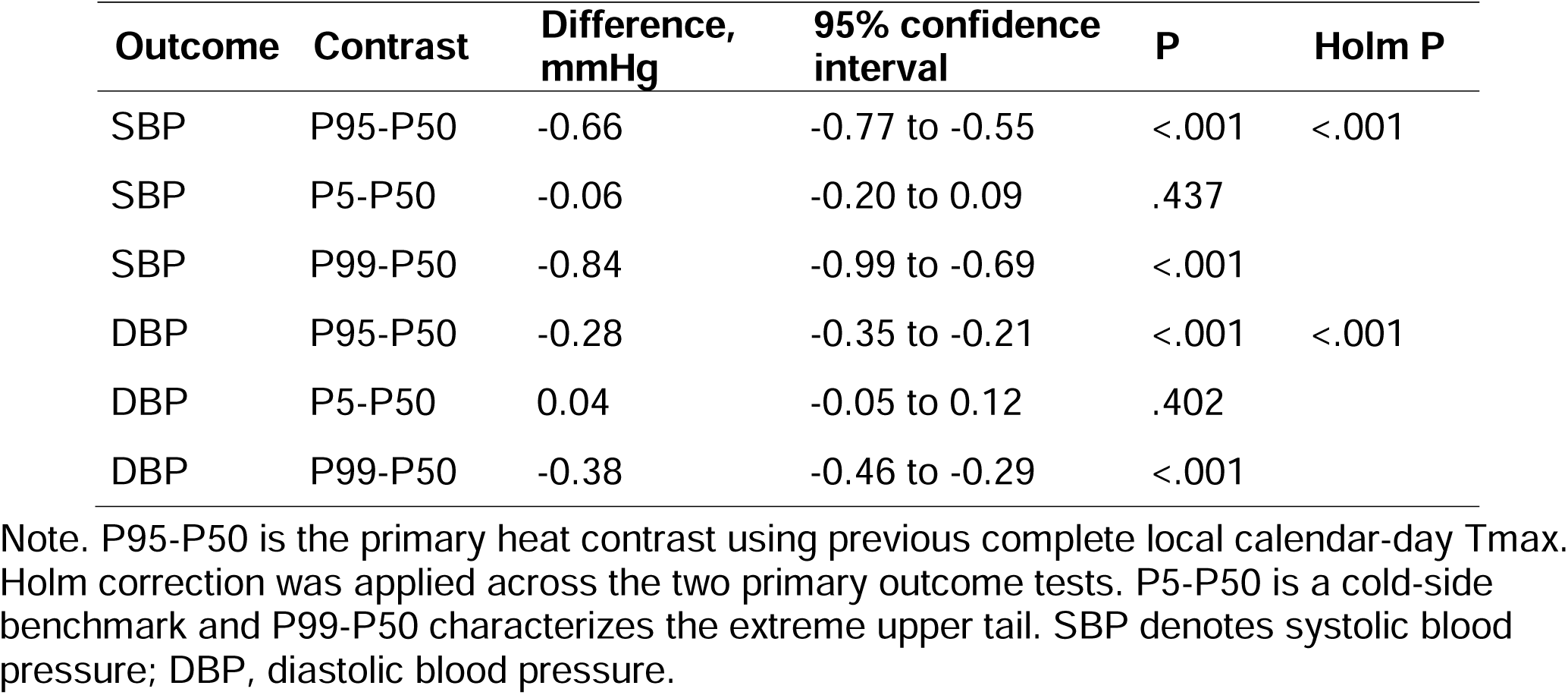
Primary and benchmark contrasts for previous complete local-day maximum temperature and blood pressure.

| **Outcome** | **Contrast** | **Difference, mmHg** | **95% confidence interval** | **P** | **Holm P** |
|---|---|---|---|---|---|
| SBP | P95-P50 | -0.66 | -0.77 to -0.55 | <.001 | <.001 |
| SBP | P5-P50 | -0.06 | -0.20 to 0.09 | .437 |  |
| SBP | P99-P50 | -0.84 | -0.99 to -0.69 | <.001 |  |
| DBP | P95-P50 | -0.28 | -0.35 to -0.21 | <.001 | <.001 |
| DBP | P5-P50 | 0.04 | -0.05 to 0.12 | .402 |  |
| DBP | P99-P50 | -0.38 | -0.46 to -0.29 | <.001 |  |

Note. P95-P50 is the primary heat contrast using previous complete local calendar-day Tmax. Holm correction was applied across the two primary outcome tests. P5-P50 is a cold-side benchmark and P99-P50 characterizes the extreme upper tail. SBP denotes systolic blood pressure; DBP, diastolic blood pressure.

**Figure 1.**
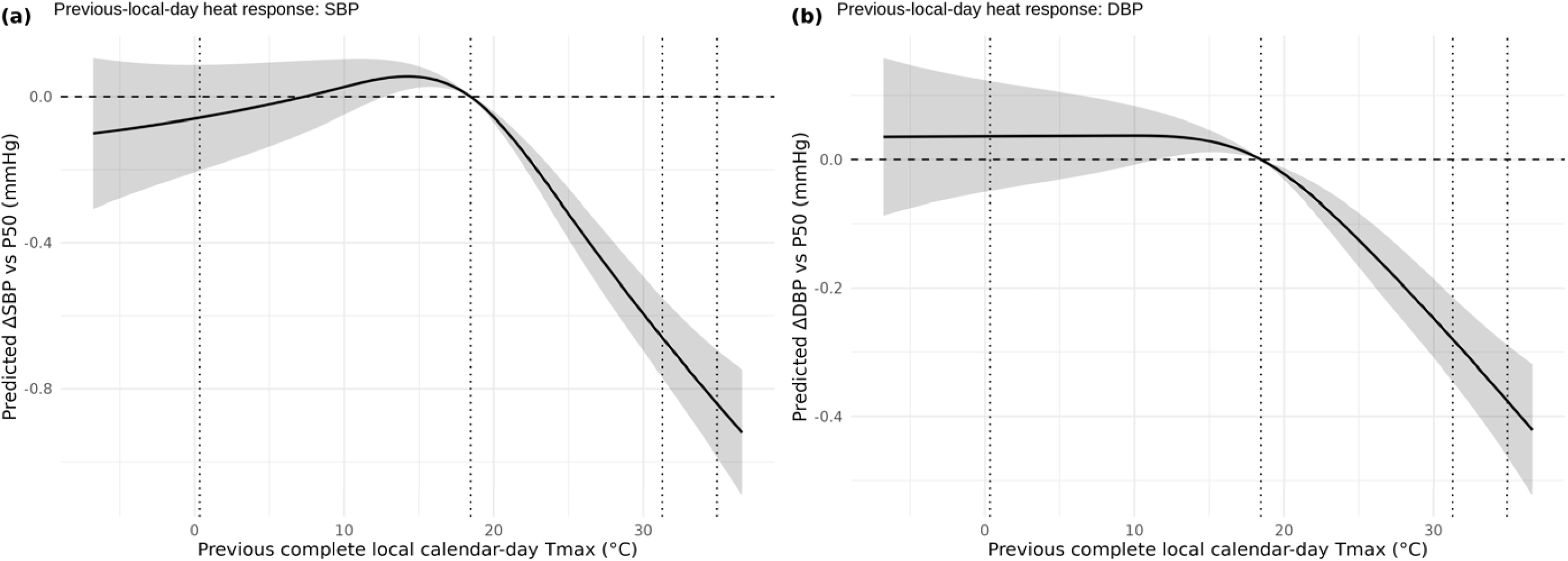
Within-person previous-local-day temperature and blood-pressure response curves. (a) Systolic blood pressure and (b) diastolic blood pressure are shown relative to the median previous complete local calendar-day maximum temperature. Models include participant and exact-date fixed effects with a common 4-degree-of-freedom natural cubic spline. Shaded bands show 95% confidence intervals. Vertical reference lines mark the 5th, 50th, 95th and 99th percentiles (P5, P50, P95 and P99). Alt text: Two panels show fitted systolic and diastolic blood-pressure differences across preceding-day maximum temperatures. Both curves decline above the median; grey bands show 95% confidence intervals.

### Age-related differences in the temperature response

Temperature-response curves varied with age in both categorical models (systolic: χ²=207.16, df=8; diastolic: χ²=219.36, df=8; both P<.001). In the categorical models, the P95-versus-P50 systolic contrasts were −0.21 mmHg among adults younger than 45 years, −0.58 mmHg at ages 45–64 and −0.98 mmHg at ages 65 years or older; the corresponding diastolic contrasts were −0.05, −0.27 and −0.42 mmHg (Supplementary Table S2). In separately fitted continuous-age models (Figure 2), systolic contrasts were −0.19 mmHg at age 35, −0.59 mmHg at age 55 and −0.99 mmHg (95% CI, −1.12 to −0.85) at age 75. Each additional decade shifted the systolic contrast by −0.20 mmHg (95% CI, −0.24 to −0.16; Holm-adjusted P<.001) and the diastolic contrast by −0.09 mmHg (95% CI, −0.12 to −0.07; Holm-adjusted P<.001). Joint continuous-age spline interactions were significant for both outcomes (systolic: χ²=232.84; diastolic: χ²=244.94; df=4 for each; both P<.001). At the colder end, the fitted diastolic curve at age 75 lay above the median-temperature reference, while the overall cold-side contrast was near zero. The pooled 95th percentile of temperature fell within the observed 1st-to-99th-percentile range of each age group.

**Figure 2.**
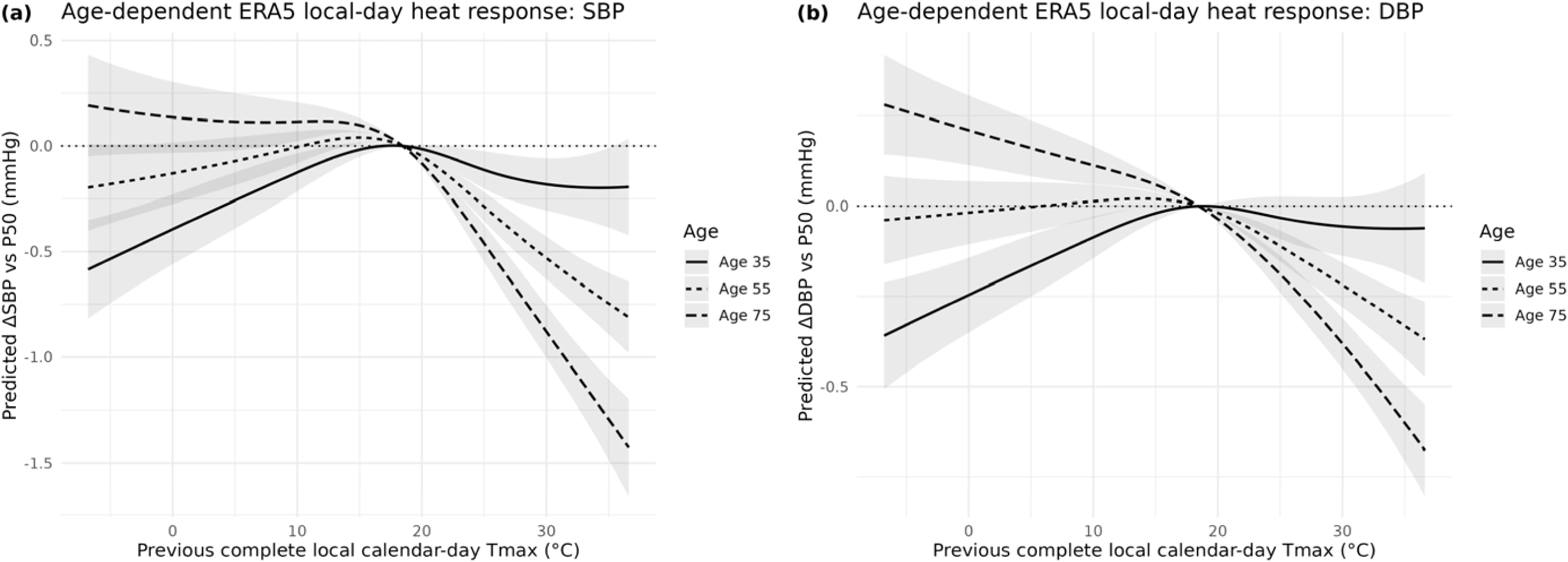
Age-dependent temperature-response curves. (a) Systolic blood pressure and (b) diastolic blood pressure are shown relative to P50 at ages 35, 55 and 75 years. At the colder end, the fitted diastolic curve at age 75 lies above the P50 reference. Curves come from the primary fixed-effects model with spline-by-continuous-age interactions; shaded bands show 95% confidence intervals. Alt text: Two panels show fitted systolic and diastolic blood-pressure responses across preceding-day temperatures at ages 35, 55 and 75. The decline at high temperatures is steeper at older ages.

### Research question 2: Social and climate context

SMV was analysed continuously, and we used its 90th-versus-10th-percentile contrast to illustrate modification of the fitted heat response. The difference in the P95-versus-P50 systolic temperature association between these SMV values was 0.21 mmHg (95% CI, −0.07 to 0.48; Holm-adjusted P=.284); the corresponding diastolic difference was −0.10 mmHg (95% CI, −0.26 to 0.06; Holm-adjusted P=.284). Joint spline-by-SMV tests showed little evidence of modification (P=.301 and P=.658, respectively; Supplementary Table S3).

Long-term cooling-degree-day normal likewise showed little evidence of modifying the acute temperature curve. Joint tests were P=.085 for systolic pressure and P=.853 for diastolic pressure (Supplementary Table S4). Fitted associations across historical climate conditions are shown in Supplementary Figure S1.

### Research question 3: Wildfire smoke and concurrent heat

Same-day wildfire-specific fine particulate matter showed small, imprecise associations with blood pressure when modelled separately (Supplementary Table S5). After joint adjustment, a 10 µg/m³ higher smoke concentration was associated with −0.03 mmHg systolic pressure (95% CI, −0.08 to 0.01; P=.124) and −0.01 mmHg diastolic pressure (95% CI, −0.04 to 0.02; P=.466). The previous-day heat contrast remained negative after smoke adjustment (−0.71 and −0.34 mmHg for systolic and diastolic pressure). Joint heat-by-smoke interaction tests yielded P=.038 for systolic pressure and P=.362 for diastolic pressure. Same-day compound-exposure estimates are also provided in Supplementary Table S5.

### Secondary outcomes and sensitivity analyses

The previous-day temperature association remained negative across alternative modelling and sample definitions (Supplementary Table S6). Averaging T_max_ over the two preceding complete days yielded systolic and diastolic P95-to-P50 contrasts of −0.49 and −0.20 mmHg. Results were similar for a three-day window and with warm-season restriction, equal-person weighting, recent residential ZIP3 restriction or ZIP3-by-year-month fixed effects. Same-day T_max_ produced a larger negative association, although this sensitivity analysis did not preserve the same temporal ordering. In the all-dates SMV sensitivity analysis, the systolic modification contrast was 0.26 mmHg (95% CI, 0.05 to 0.48; P=.017), whereas the diastolic estimate was −0.01 mmHg (95% CI, −0.14 to 0.12; P=.899). Pulse pressure and mean arterial pressure also declined, and spline comparisons retained a negative extreme-tail association (Supplementary Table S7).

For visit-to-visit blood-pressure variability, the overall P95-to-P50 ASD contrast was 0.02 mmHg for systolic pressure (95% CI, −0.07 to 0.11; Holm-adjusted P=.661) and 0.05 mmHg for diastolic pressure (95% CI, −0.01 to 0.11; Holm-adjusted P=.162). However, the systolic ASD contrast became 0.05 mmHg more positive per decade of age (95% CI, 0.01 to 0.08; Holm-adjusted P=.009; joint interaction P=.020). The diastolic age interaction was weaker. SMV modification was not clearly supported, and shorter interval sensitivity results were similar (Supplementary Table S8; Supplementary Figure S2).

## Discussion

In outcome-specific panels exceeding 2.2 million outpatient person-days, warmer previous-day temperatures were associated with lower systolic and diastolic blood pressure within individuals. The continuous association extended through the extreme upper tail, and the temperature contrast became progressively more negative with age. Together, the findings add population-based evidence of age-related differences in blood-pressure responses to heat.

The results were broadly consistent with those reported by McGrath et al. (2026) in a separate US outpatient population. That study found approximately −0.7 mmHg systolic and −0.3 mmHg diastolic differences during same-day extreme heat, with a −1.1 mmHg systolic difference among adults aged 65 years or older. Our corresponding estimates were −0.66 and −0.28 mmHg, and the systolic contrast reached −0.99 mmHg at age 75. The studies used different temperature contrasts and exposure windows. Our continuous temperature model further describes the upper-tail curve, while participant and exact calendar-date effects identify the association from within-person changes. The preceding complete local-day window provides a defined measurement sequence.

The previous-local-day definition ensured that the temperature window ended before the date of clinical measurement. This choice also aligns with McGrath et al.’s (2026) lag analysis, in which the strongest differences occurred on the exposure day and following day. We observed negative associations using mean temperatures from the preceding two and three complete days. Same-day Tmax gave a larger estimate but could include temperature measured after the clinical reading.

The inverse association persisted at the extreme upper tail. Blood pressure at the 99th temperature percentile remained lower than at the 95th in the spline analyses. During heat stress, redistribution of blood toward the skin lowers systemic vascular resistance, while sweating and volume loss challenge venous return. Laboratory studies also document changes in stroke volume and ventricular filling (Foster et al., 2025; Meade et al., 2025).

These mechanisms offer a plausible explanation for the observed average-pressure pattern. Higher pressure during heatwave warnings in a separate study of older hypertensive adults (Zhang et al., 2026) illustrates that responses during acute heat emergencies may differ from associations observed across routine outpatient visits.

The clearest modification was by age: the high-versus-median temperature difference became progressively more negative for systolic pressure across adulthood. This may reflect age-related physiological challenges during heat exposure, including reduced capacity to preserve central blood volume (Cottle et al., 2025). Older adults often use medicines that affect vascular tone or fluid balance. At the colder end, the fitted diastolic curve at age 75 was above its median-temperature reference (Figure 2), despite a near-zero overall cold-side contrast. This direction resembles findings from the Three-City Study of older adults, where colder outdoor temperatures were found to be associated with higher blood pressure (Alpérovitch et al., 2009). The Three-City Study included only adults aged 65 years or older and found larger longitudinal temperature associations among those aged 80 years or older. The broader age range in our cohort may partly explain the weaker overall cold-side association. Cold-related mortality remains a public health concern (Paniello-Castillo et al., 2026).

The secondary analysis suggested little overall relation between heat and absolute changes in blood pressure across successive outpatient visits. The temperature contrast in systolic ASD increased modestly with age. As these intervals are irregular and can span months, they describe a different aspect of variation from short-term ambulatory fluctuations (Supplementary Table S8).

The high-temperature association showed little evidence of modification by SMV or historical climate context. It also remained negative after adjustment for wildfire-specific particulate matter, although the spline-by-smoke interaction test suggested heterogeneity for systolic pressure in the smoke-covered period. These contextual analyses support the consistency of the primary inverse temperature association, while leaving the age gradient as the main source of heterogeneity.

The magnitude of the association is important for interpretation. In descriptive analyses, median within-person visit-to-visit standard deviations were 10.62 mmHg for systolic and 6.89 mmHg for diastolic pressure (Supplementary Table S8), substantially larger than the mean P95-versus-P50 temperature contrasts of −0.66 and −0.28 mmHg. These descriptive quantities arise from related but different analytic samples. The average differences are therefore unlikely to alter treatment decisions at an individual visit. The consistency of the association across cohorts and its age gradient inform our understanding of cardiovascular responses to heat.

The national repeated-measures design offered substantial within-person information, with several limitations. Residential ZIP3 was generalised and could not capture indoor temperature or time spent away from home. EHR measurements arose during routine visits, without standardised information on medication timing, hydration or transient symptoms. Heat may also influence who seeks care. All of Us is a non-probability sample, and repeat measurements occur more often among people engaged in healthcare. Survey availability reduced the SMV sample, and the composite did not directly measure household cooling access. Wildfire-smoke data ended in 2023, shortening the observation window and reducing the sample available for interaction analyses. BP variability was based on absolute differences across irregular visit intervals rather than ambulatory monitoring. Drug-specific interactions and direct haemodynamic mechanisms were not evaluated. Time-varying confounding remains possible, and these associations do not establish causality.

## Conclusion

In this large US outpatient cohort, encompassing more than 2.2 million outcome-specific person-days, higher temperature during the preceding complete local day was associated with modestly lower blood pressure within individuals. The inverse association persisted into the extreme upper tail and became progressively stronger with age. These findings describe an age-dependent component of cardiovascular responses to ambient heat in routine outpatient care.

## Acknowledgments

We gratefully acknowledge All of Us participants for their contributions, without whom this research would not have been possible. We also thank the National Institutes of Health’s All of Us Research Program for making available the participant data examined in this study.

## AI disclosure

ChatGPT (OpenAI) assisted with language editing. The authors directed the work, reviewed the outputs and are responsible for the accuracy and integrity of the manuscript.

## Author contributions

SY conceptualised the study with DAS, YCK and HEB. SY curated the data, wrote the analysis software, performed the formal analyses, produced the visualisations and drafted the manuscript. All authors contributed to methodology and manuscript review and editing. DAS, YCK and HEB provided supervision. SY had full access to all study data and takes responsibility for the integrity and accuracy of the reporting.

## Data availability

This study used data from the All of Us Research Program Controlled Tier Dataset C2025Q4R6, available to authorised users on the Researcher Workbench under program data-use requirements. Acute meteorology came from the Copernicus Climate Change Service ERA5-Land dataset. Long-term climate normals came from the PRISM Climate Group, and wildfire-smoke estimates came from the Stanford ECHO resource. U.S. Census and American Community Survey data supported geographic aggregation where required. The analysis-ready public environmental data can be regenerated from the accompanying processing code.

## Code availability

Code for the All of Us analysis pipeline will be deposited in a public repository at publication.

## Ethics

The All of Us Research Program operates under centralised institutional review board oversight, and participants provide informed consent for use of their data in approved research. This study used de-identified Controlled Tier data within the secure Researcher Workbench and followed the All of Us Data Use Agreement.

## Participant and public involvement

This secondary analysis did not include additional study-specific participant or public involvement beyond the All of Us Research Program’s participant-partner framework.

