## Supplementary Material for "Ambient Heat and Age-dependent Blood Pressure Responses in 184,000 U.S. Adults"

**Supplementary Methods**

**ERA5-Land acute meteorology and local-day construction**

Acute meteorology was obtained from the official ERA5-Land hourly Analysis Ready Cloud Optimised Zarr archive (Muñoz-Sabater et al., 2021). The extraction used 2-m air temperature, 2-m dew-point temperature and total precipitation. A fixed mapping linked each generalized three-digit ZIP-code prefix (ZIP3) to an ERA5-Land grid cell and a representative-point Internet Assigned Numbers Authority time zone. Selected physical grid cells were downloaded once and reused when the same cell contributed to more than one ZIP3. Hourly timestamps were converted from Coordinated Universal Time to the mapped local time zone before daily aggregation. Hourly air temperature yielded daily maximum temperature (Tmax), mean temperature (Tmean) and minimum temperature (Tmin). Dew point was averaged within each local day, and hourly precipitation was summed. Daily completeness was evaluated against the expected number of local hours, including 23-hour and 25-hour daylight-saving days. Extraction began October 1, 2021 so lagged exposures were available before the analytic start date. The resulting local-day dataset contained 881 ZIP3s and 1,046,628 ZIP3-day rows through December 31, 2024. The primary exposure was previous complete local calendar-day Tmax (local t-1). A secondary exposure averaged Tmax over local t-1 and t-2.

**Blood-pressure construction**

Clinical electronic health record (EHR) blood-pressure measurements came from the All of Us Controlled Tier Curated Data Repository version 9 Observational Medical Outcomes Partnership measurement table. All of Us enrollment physical-measurement source concepts were excluded. Primary analyses used outpatient or ambulatory encounters. Systolic blood pressure (SBP) outside 70-250 mmHg and diastolic blood pressure (DBP) outside 40-150 mmHg were removed. Multiple same-day readings of the same type were averaged within participant and setting. Pulse pressure and mean arterial pressure were derived when both daily values were available and SBP exceeded DBP. Measurement date and measurement_datetime were retained for timing diagnostics. Outpatient measurements had a median hour of 12:00 (interquartile range [IQR], 09:00-15:00); 45.2% occurred before noon and 71.7% before 15:00.

**Social-material vulnerability construction**

The social-material vulnerability (SMV) index date was the first Social Determinants of Health survey date on or after November 1, 2021 with scoreable loneliness and functional rurality or resource-access domains. The survey-based components captured loneliness and resource access, then added social support and neighborhood social context. Material hardship represented immediate resource strain. Income, education and ZIP3 deprivation supplied broader socioeconomic and area context. Survey items used question-specific response mappings, and protective items were reverse-coded so higher scores indicated greater vulnerability. Loneliness required at least 4 of 8 items and functional rurality required at least 3 of 5. Social support required at least 4 of 8 items, while neighborhood cohesion required at least 3 of 5. Neighborhood disorder required at least 7 of 14 items and material hardship required at least 2 of 3. Income and education used the latest substantive response on or before the index date. The area-level deprivation index was obtained from the CDR v9 zip3_ses_map table (deprivation_index), which contains ZIP3-linked American Community Survey socioeconomic data (All of Us Research Program, 2026; Brokamp et al., 2019). The index reflects six area-level measures of material deprivation, including poverty, income and educational attainment. Higher values indicate greater deprivation. We normalised the ZIP3 identifier, averaged available deprivation values if multiple records mapped to the same ZIP3 and assigned the index using the most recent generalised residential ZIP3 observed on or before the survey index date. Retained components were standardized separately. Participant-level SMV was the equal-weight mean of available component z scores when at least three components were observed, followed by a second standardization.

**Demographic heterogeneity and continuous-age analysis**

Age heterogeneity used age at each blood-pressure measurement, classified as younger than 45 years, 45–64 years or 65 years or older. Categorical-age models included the corresponding time-varying group main effects and spline-by-group interactions, alongside participant and calendar-date fixed effects. During follow-up, 13,983 of 181,865 participants (7.69%) in the systolic model and 13,967 of 181,770 (7.68%) in the diastolic model contributed observations in more than one age group. Group-specific participant counts can therefore overlap. The separately fitted continuous-age model interacted the common four-degree-of-freedom temperature spline with age at each measurement; estimates at ages 35, 55 and 75 years and the change per decade were obtained from that model. Joint Wald tests assessed eight spline-by-group interaction terms in the categorical model and four spline-by-age terms in the continuous-age model. Common-support diagnostics compared the pooled P95 with each age group's P1–P99 temperature distribution. Gender and race grouping followed the Curated Data Repository concepts; race was defined separately from ethnicity.

**Wildfire-smoke processing**

Wildfire-specific fine particulate matter with aerodynamic diameter 2.5 µm or smaller (PM2.5) came from Stanford Environmental Change and Human Outcomes (ECHO) estimates based on the Childs et al. framework (Childs et al., 2022). ZIP Code Tabulation Area (ZCTA)-day values were population-weighted to ZIP3. Within verified coverage, ZCTA-days without a smoke record contributed zero wildfire-attributable PM2.5 while remaining in the ZIP3 population denominator. Dates outside verified coverage remained missing. Smoke analysis therefore ended on December 31, 2023. Smoke-only models adjusted for a 3-degree-of-freedom spline of same-day ERA5-Land mean temperature with same-day dew point and precipitation. Mutually adjusted models combined same-day smoke with the previous-local-day heat spline.

**Blood-pressure variability and quality-control framework**

Participant-level blood-pressure variability (BPV) descriptors included standard deviation (SD), coefficient of variation (CV) and average real variability (ARV) among participants with at least three eligible days. Inferential models used absolute successive difference (ASD), defined as the absolute change from the immediately preceding outcome-specific blood-pressure value. Intervals longer than 365 days were excluded from the primary BPV analysis. Models adjusted for log1p interval length in days, and participants required at least two usable intervals. A 90-day maximum interval was used in sensitivity analysis. For the post-index SMV analysis, intervals were constructed after restricting the panel to measurements on or after the SMV index date, so both contributing measurements were post-index. The ERA5-Land ZIP3-day dataset was checked for duplicate ZIP3-date records, complete daily coverage, expected local-day hour counts and plausible meteorologic values. Spline construction was verified before model fitting. Analyses used R 4.5.3 and fixest 0.14.2. The complete primary mean-pressure model included four fixed natural-spline basis terms for previous-day Tmax, participant and exact calendar-date fixed effects, previous-day mean dew point and precipitation. P95–P50 contrasts were obtained from the difference between spline basis vectors evaluated at the two temperature anchors, with standard errors computed using the participant-clustered coefficient covariance matrix. No temperature percentile groups were created.

**Supplementary Table S1. Environmental, blood-pressure and analytic-panel quality control.**

| **Domain** | **Metric** | **Value** |
| --- | --- | --- |
| ERA5-Land local-day dataset | ZIP3-day rows | 1,046,628 |
| ERA5-Land local-day dataset | ZIP3s | 881 |
| ERA5-Land local-day dataset | Date range | 2021-10-01 to 2024-12-31 |
| ERA5-Land local-day dataset | Duplicate ZIP3-date rows | 0 |
| Primary exposure | Tmax P50 / P95 / P99 | 18.45 / 31.28 / 34.92 °C |
| Outpatient BP | Person-days / persons | 2,448,649 / 219,251 |
| ZIP3 + ERA5 linked | Person-days / persons | 2,372,695 / 212,693 |
| Primary weather-linked | Person-days / persons | 2,367,677 / 212,135 |
| Primary SBP model | Person-days / persons | 2,279,955 / 181,865 |
| Primary DBP model | Person-days / persons | 2,266,080 / 181,770 |
| Post-index SMV weather-linked panel | Person-days / persons | 866,244 / 84,548 |
| Post-index SMV SBP model | Person-days / persons | 844,056 / 82,932 |
| Post-index SMV DBP model | Person-days / persons | 839,478 / 82,795 |
| Smoke verified ECHO coverage | Person-days / persons | 1,678,511 / 200,567 |
| Smoke SBP model | Person-days / persons | 1,598,983 / 166,729 |
| Smoke DBP model | Person-days / persons | 1,595,110 / 166,613 |
| Primary BPV SBP | Intervals / persons | 2,034,434 / 153,359 |
| Primary BPV DBP | Intervals / persons | 2,020,644 / 153,286 |
| Post-index SMV BPV SBP | Intervals / persons | 735,799 / 67,438 |
| Post-index SMV BPV DBP | Intervals / persons | 731,541 / 67,463 |
| Outpatient BP timing | Measurement hour | Median 12:00 [IQR, 09:00-15:00]; 45.2% before noon; 71.7% before 15:00 |

Note. BP denotes blood pressure; SBP, systolic blood pressure; DBP, diastolic blood pressure; BPV, blood-pressure variability; ECHO, Environmental Change and Human Outcomes. Primary-analysis weather-linked counts precede outcome-specific repeated-measure and complete-case filtering. BPV intervals are absolute successive differences with a maximum 365-day interval.

**Supplementary Table S2. Demographic heterogeneity in the P95-versus-P50 previous-local-day temperature contrast.**

| **Modifier** | **Outcome** | **Group/test** | **Estimate / support** | **P** | **Detail** |
| --- | --- | --- | --- | --- | --- |
| Age | SBP | <45 | -0.21 (-0.35 to -0.06) | .004 | n=51,813 |
| Age | DBP | <45 | -0.05 (-0.14 to 0.05) | .350 | n=51,798 |
| Age | SBP | 45-64 | -0.58 (-0.72 to -0.44) | <.001 | n=74,301 |
| Age | DBP | 45-64 | -0.27 (-0.36 to -0.19) | <.001 | n=74,240 |
| Age | SBP | 65+ | -0.98 (-1.12 to -0.84) | <.001 | n=69,734 |
| Age | DBP | 65+ | -0.42 (-0.50 to -0.34) | <.001 | n=69,699 |
| Age | SBP | Joint curve heterogeneity | χ²=207.16, df=8 | <.001 |  |
| Age | DBP | Joint curve heterogeneity | χ²=219.36, df=8 | <.001 |  |
| Gender | SBP | Female | -0.57 (-0.68 to -0.45) | <.001 | n=119,889 |
| Gender | DBP | Female | -0.23 (-0.31 to -0.16) | <.001 | n=119,812 |
| Gender | SBP | Male | -0.88 (-1.04 to -0.73) | <.001 | n=59,220 |
| Gender | DBP | Male | -0.38 (-0.47 to -0.29) | <.001 | n=59,190 |
| Gender | SBP | Other/Unknown | -0.34 (-0.89 to 0.22) | .235 | n=2,756 |
| Gender | DBP | Other/Unknown | -0.25 (-0.55 to 0.06) | .117 | n=2,768 |
| Gender | SBP | Joint curve heterogeneity | χ²=91.11, df=8 | <.001 |  |
| Gender | DBP | Joint curve heterogeneity | χ²=65.10, df=8 | <.001 |  |
| Race | SBP | White | -0.64 (-0.76 to -0.51) | <.001 | n=117,872 |
| Race | DBP | White | -0.32 (-0.40 to -0.24) | <.001 | n=117,803 |
| Race | SBP | Black | -0.61 (-0.82 to -0.41) | <.001 | n=27,736 |
| Race | DBP | Black | -0.34 (-0.45 to -0.23) | <.001 | n=27,720 |
| Race | SBP | Asian | -0.84 (-1.24 to -0.45) | <.001 | n=5,374 |
| Race | DBP | Asian | -0.22 (-0.47 to 0.03) | .086 | n=5,376 |
| Race | SBP | Multiple race | -0.64 (-1.09 to -0.18) | .006 | n=3,305 |
| Race | DBP | Multiple race | -0.25 (-0.54 to 0.04) | .089 | n=3,305 |
| Race | SBP | Other | -1.30 (-1.90 to -0.70) | <.001 | n=1,765 |
| Race | DBP | Other | -0.59 (-0.95 to -0.23) | .001 | n=1,759 |
| Race | SBP | Joint curve heterogeneity | χ²=30.99, df=16 | .013 |  |
| Race | DBP | Joint curve heterogeneity | χ²=24.81, df=16 | .073 |  |
| Continuous age | SBP | Age 35 | -0.19 (-0.32 to -0.06) | .005 |  |
| Continuous age | SBP | Age 55 | -0.59 (-0.70 to -0.48) | <.001 |  |
| Continuous age | SBP | Age 75 | -0.99 (-1.12 to -0.85) | <.001 |  |
| Continuous age | SBP | Per +10 years | -0.20 (-0.24 to -0.16) | <.001 | Holm P<.001 |
| Continuous age | SBP | Joint curve heterogeneity | χ²=232.84, df=4 | <.001 | n=181,865 |
| Continuous age | DBP | Age 35 | -0.06 (-0.15 to 0.03) | .182 |  |
| Continuous age | DBP | Age 55 | -0.25 (-0.31 to -0.18) | <.001 |  |
| Continuous age | DBP | Age 75 | -0.44 (-0.51 to -0.36) | <.001 |  |
| Continuous age | DBP | Per +10 years | -0.09 (-0.12 to -0.07) | <.001 | Holm P<.001 |
| Continuous age | DBP | Joint curve heterogeneity | χ²=244.94, df=4 | <.001 | n=181,770 |
| Age temperature support | All | <45 | P1-P99: -6.97 to 35.00 °C |  | n=62,875; 23.6% with ≥1 day at/above pooled P95 |
| Age temperature support | All | 45-64 | P1-P99: -6.47 to 34.91 °C |  | n=85,332; 26.1% with ≥1 day at/above pooled P95 |
| Age temperature support | All | 65+ | P1-P99: -6.83 to 34.90 °C |  | n=78,145; 27.4% with ≥1 day at/above pooled P95 |

Note. Categorical age was defined at each measurement; the categorical models included age-group main effects and spline-by-age-group interactions, with participant and calendar-date fixed effects. In the systolic and diastolic models, 7.69% and 7.68% of participants, respectively, contributed measurements in more than one age group, so group-specific counts are not mutually exclusive. The categorical-age estimates shown here use the specification retaining identifiable group main effects. Continuous-age models were fitted separately using the same primary temperature spline. Per-decade estimates quantify changes in the P95-versus-P50 contrast with age. Joint P values test all spline-by-modifier interactions. Gender and race categories came from the Curated Data Repository; race grouping did not incorporate ethnicity.

**Supplementary Table S3. Social-material vulnerability components, common support and primary modification tests.**

| **Section** | **Item/outcome** | **Value / estimate** | **Decision / P** |
| --- | --- | --- | --- |
| Component | Loneliness | 0.0% missing | Retained |
| Component | Functional rurality / resource access | 0.0% missing | Retained |
| Component | Low social support | 0.6% missing | Retained |
| Component | Low neighborhood cohesion | 1.4% missing | Retained |
| Component | Neighborhood disorder | 0.5% missing | Retained |
| Component | Material hardship | 0.4% missing | Retained |
| Component | Low income | 10.8% missing | Retained |
| Component | Low education | 1.4% missing | Retained |
| Component | ZIP3 deprivation | 3.9% missing | Retained |
| Common support | SMV P10 | -1.267 |  |
| Common support | SMV P50 | -0.279 |  |
| Common support | SMV P90 | 1.144 |  |
| Common support | Low SMV quintile temp P1 | -5.991 °C |  |
| Common support | Low SMV quintile temp P99 | 34.535 °C |  |
| Common support | High SMV quintile temp P1 | -5.874 °C |  |
| Common support | High SMV quintile temp P99 | 35.132 °C |  |
| Common support | Pooled temp P50 | 18.649 °C |  |
| Common support | Pooled temp P95 | 31.132 °C |  |
| Common support | Selected high temperature | 31.132 °C |  |
| Common support | Selected rule | P95 |  |
| Primary SMV modification | SBP | 0.21 (-0.07 to 0.48) | Holm P=.284 |
| Joint spline × SMV | SBP | χ²=4.87, df=4 | P=.301 |
| Primary SMV modification | DBP | -0.10 (-0.26 to 0.06) | Holm P=.284 |
| Joint spline × SMV | DBP | χ²=2.43, df=4 | P=.658 |

Note. The post-index SMV analysis contrast is the difference in the P95-to-P50 heat response between SMV P90 and P10. P95 satisfied the common-support rule. Component missingness is calculated in the SMV construction sample (N=302,710).

**Supplementary Table S4. Long-term cooling-degree-day (CDD) climate context and spline-heterogeneity tests.**

| **Section** | **Percentile/outcome** | **CDD normal or statistic** | **Standardized value / P** |
| --- | --- | --- | --- |
| CDD distribution | P10 | 197.25 | z=-0.692 |
| CDD distribution | P50 | 374.56 | z=-0.299 |
| CDD distribution | P90 | 1036.66 | z=1.172 |
| Joint spline × CDD | SBP | χ²=8.20, df=4 | P=.085 |
| Joint spline × CDD | DBP | χ²=1.35, df=4 | P=.853 |

Note. Higher cooling-degree-day (CDD) normal indicates a historically warmer climate. CDD was derived from 1991-2020 normals from the Parameter-elevation Regressions on Independent Slopes Model (PRISM) and was used only as long-term climate context. Acute heat came from ERA5-Land local-day meteorology. Model-implied curves are shown in Supplementary Figure S1.

**Supplementary Table S5. Wildfire-smoke and compound heat-smoke analyses.**

| **Analysis** | **Outcome** | **Contrast** | **Estimate / statistic** | **P** |
| --- | --- | --- | --- | --- |
| Smoke-only | SBP | +10 µg/m³ ECHO wildfire PM2.5; adjusted for same-local-day ERA5 Tmean | 0.03 (-0.02 to 0.07) | .259 |
| Mutually adjusted | SBP | +10 µg/m³ smoke adjusted for previous-local-day nonlinear heat | -0.03 (-0.08 to 0.01) | .124 |
| Mutually adjusted | SBP | Previous-local-day heat P95-P50 adjusted for same-day smoke | -0.71 (-0.84 to -0.58) | <.001 |
| Same-local-day compound categories | SBP | Heat only | -0.64 (-0.88 to -0.40) | <.001 |
| Same-local-day compound categories | SBP | Smoke only | -0.03 (-0.10 to 0.04) | .433 |
| Same-local-day compound categories | SBP | Heat + smoke | -0.28 (-0.42 to -0.14) | <.001 |
| Smoke-only | DBP | +10 µg/m³ ECHO wildfire PM2.5; adjusted for same-local-day ERA5 Tmean | 0.01 (-0.02 to 0.04) | .441 |
| Mutually adjusted | DBP | +10 µg/m³ smoke adjusted for previous-local-day nonlinear heat | -0.01 (-0.04 to 0.02) | .466 |
| Mutually adjusted | DBP | Previous-local-day heat P95-P50 adjusted for same-day smoke | -0.34 (-0.42 to -0.26) | <.001 |
| Same-local-day compound categories | DBP | Heat only | -0.35 (-0.49 to -0.20) | <.001 |
| Same-local-day compound categories | DBP | Smoke only | -0.05 (-0.09 to -0.00) | .032 |
| Same-local-day compound categories | DBP | Heat + smoke | -0.12 (-0.20 to -0.03) | .007 |
| Heat × smoke | SBP | Joint nonlinear heat × smoke | χ²=10.16, df=4 | .038 |
| Heat × smoke | DBP | Joint nonlinear heat × smoke | χ²=4.34, df=4 | .362 |

Note. The smoke analysis ended on December 31, 2023. Smoke-only models adjust for same-day ERA5-Land mean temperature, dew point and precipitation. Mutually adjusted models combine same-day smoke with previous-local-day heat. Compound categories use neither exposure as the reference.

**Supplementary Table S6. Sensitivity and robustness analyses of previous-local-day temperature exposure.**

| **Analysis** | **Outcome** | **Exposure** | **Contrast estimate, mmHg (95% confidence interval)** | **P** |
| --- | --- | --- | --- | --- |
| Alternative ERA5 previous-local-day Tmax, spline df=3 | SBP | Previous-day Tmax | -0.58 (-0.68 to -0.47) | <.001 |
| Alternative ERA5 previous-local-day Tmax, spline df=3 | DBP | Previous-day Tmax | -0.26 (-0.32 to -0.20) | <.001 |
| Alternative ERA5 previous-local-day Tmax, spline df=5 | SBP | Previous-day Tmax | -0.64 (-0.75 to -0.53) | <.001 |
| Alternative ERA5 previous-local-day Tmax, spline df=5 | DBP | Previous-day Tmax | -0.29 (-0.36 to -0.23) | <.001 |
| Secondary: mean Tmax over two complete local days preceding BP (t-1, t-2) | SBP | Previous two-day mean Tmax | -0.49 (-0.61 to -0.37) | <.001 |
| Secondary: mean Tmax over two complete local days preceding BP (t-1, t-2) | DBP | Previous two-day mean Tmax | -0.20 (-0.27 to -0.12) | <.001 |
| Sensitivity: same local calendar-day Tmax (not strictly temporally ordered) | SBP | Same-day Tmax | -0.89 (-1.00 to -0.78) | <.001 |
| Sensitivity: same local calendar-day Tmax (not strictly temporally ordered) | DBP | Same-day Tmax | -0.33 (-0.40 to -0.27) | <.001 |
| Sensitivity: mean Tmax over three complete local days preceding BP (t-1:t-3) | SBP | Previous three-day mean Tmax | -0.40 (-0.53 to -0.27) | <.001 |
| Sensitivity: mean Tmax over three complete local days preceding BP (t-1:t-3) | DBP | Previous three-day mean Tmax | -0.15 (-0.23 to -0.08) | <.001 |
| Previous complete local-day Tmean | SBP | Previous-day Tmean | -0.65 (-0.80 to -0.50) | <.001 |
| Previous complete local-day Tmean | DBP | Previous-day Tmean | -0.29 (-0.38 to -0.20) | <.001 |
| Previous complete local-day Tmin | SBP | Previous-day Tmin | -0.25 (-0.41 to -0.09) | .002 |
| Previous complete local-day Tmin | DBP | Previous-day Tmin | -0.09 (-0.18 to 0.01) | .066 |
| At least 3 outcome-specific BP days | SBP | Previous-day Tmax | -0.66 (-0.77 to -0.55) | <.001 |
| At least 3 outcome-specific BP days | DBP | Previous-day Tmax | -0.28 (-0.34 to -0.21) | <.001 |
| At least 5 outcome-specific BP days | SBP | Previous-day Tmax | -0.68 (-0.79 to -0.57) | <.001 |
| At least 5 outcome-specific BP days | DBP | Previous-day Tmax | -0.27 (-0.34 to -0.20) | <.001 |
| Equal total weight per participant | SBP | Previous-day Tmax | -0.56 (-0.70 to -0.42) | <.001 |
| Equal total weight per participant | DBP | Previous-day Tmax | -0.29 (-0.38 to -0.20) | <.001 |
| May-September only | SBP | Previous-day Tmax | -0.67 (-0.84 to -0.50) | <.001 |
| May-September only | DBP | Previous-day Tmax | -0.35 (-0.45 to -0.25) | <.001 |
| BP date within 365 days of generalized ZIP3 observation | SBP | Previous-day Tmax | -0.64 (-0.83 to -0.45) | <.001 |
| BP date within 365 days of generalized ZIP3 observation | DBP | Previous-day Tmax | -0.33 (-0.45 to -0.21) | <.001 |
| Participant + date + ZIP3 × year-month fixed effects | SBP | Previous-day Tmax | -0.71 (-0.84 to -0.59) | <.001 |
| Participant + date + ZIP3 × year-month fixed effects | DBP | Previous-day Tmax | -0.33 (-0.41 to -0.25) | <.001 |
| Multiway cluster: participant + ZIP3-year-month | SBP | Previous-day Tmax | -0.66 (-0.78 to -0.54) | <.001 |
| Multiway cluster: participant + ZIP3-year-month | DBP | Previous-day Tmax | -0.28 (-0.36 to -0.20) | <.001 |
| SMV sensitivity: all eligible BP dates regardless of SMV index date | SBP | Previous-day Tmax | 0.26 (0.05 to 0.48) | .017 |
| SMV sensitivity: all eligible BP dates regardless of SMV index date | DBP | Previous-day Tmax | -0.01 (-0.14 to 0.12) | .899 |

Note. Rows using the primary exposure report contrasts at the primary P50 and P95 unless an analysis-specific exposure distribution is indicated. Same-day Tmax is a temporally non-strict sensitivity. The sensitivity analysis for SMV modification used all eligible blood-pressure dates regardless of the SMV index date.

**Supplementary Table S7. Secondary pulse pressure (PP), mean arterial pressure (MAP) and right-tail shape diagnostics.**

| **Analysis** | **Outcome** | **Contrast** | **Estimate, mmHg (95% confidence interval)** | **P** |
| --- | --- | --- | --- | --- |
| Secondary BP phenotype | PP | P95-P50 | -0.40 (-0.49 to -0.31) | <.001 |
| Secondary BP phenotype | PP | P5-P50 | -0.08 (-0.19 to 0.04) | .182 |
| Secondary BP phenotype | PP | P99-P50 | -0.50 (-0.62 to -0.38) | <.001 |
| Secondary BP phenotype | MAP | P95-P50 | -0.42 (-0.49 to -0.35) | <.001 |
| Secondary BP phenotype | MAP | P5-P50 | -0.01 (-0.10 to 0.09) | .897 |
| Secondary BP phenotype | MAP | P99-P50 | -0.56 (-0.65 to -0.46) | <.001 |
| Right-tail diagnostic | SBP | df=3; P99-P95 | -0.26 (-0.31 to -0.20) | <.001 |
| Right-tail diagnostic | DBP | df=3; P99-P95 | -0.12 (-0.15 to -0.09) | <.001 |
| Right-tail diagnostic | SBP | df=4; P99-P95 | -0.18 (-0.25 to -0.11) | <.001 |
| Right-tail diagnostic | DBP | df=4; P99-P95 | -0.10 (-0.14 to -0.06) | <.001 |
| Right-tail diagnostic | SBP | df=5; P99-P95 | -0.19 (-0.27 to -0.12) | <.001 |
| Right-tail diagnostic | DBP | df=5; P99-P95 | -0.08 (-0.12 to -0.03) | <.001 |

Note. Pulse pressure and mean arterial pressure are derived only from physiologically ordered daily systolic-diastolic pairs. Negative P99-to-P95 estimates indicate continued decline across the extreme upper tail.

**Supplementary Table S8. Secondary visit-to-visit blood-pressure variability analyses.**

| **Section** | **Outcome/test** | **Estimate / statistic** | **P** | **Sample / detail** |
| --- | --- | --- | --- | --- |
| Participant-level descriptive | SBP | Median SD 10.62; CV 8.5%; ARV 11.60 |  | n=161,012; median 9 BP days |
| Participant-level descriptive | DBP | Median SD 6.89; CV 9.2%; ARV 7.50 |  | n=160,919; median 9 BP days |
| Primary overall heat → ASD | SBP | 0.02 (-0.07 to 0.11) | Holm P=.661 | n=153,359; intervals=2,034,434 |
| Primary ≤90-day interval sensitivity | SBP | 0.03 (-0.07 to 0.13) | .588 | n=127,438 |
| Primary overall heat → ASD | DBP | 0.05 (-0.01 to 0.11) | Holm P=.162 | n=153,286; intervals=2,020,644 |
| Primary ≤90-day interval sensitivity | DBP | 0.05 (-0.01 to 0.12) | .103 | n=127,406 |
| Primary age modification | SBP, age 35 | -0.09 (-0.20 to 0.02) | .105 | P95-P50 ASD contrast |
| Primary age modification | SBP, age 55 | 0.00 (-0.09 to 0.09) | .991 | P95-P50 ASD contrast |
| Primary age modification | SBP, age 75 | 0.09 (-0.02 to 0.20) | .093 | P95-P50 ASD contrast |
| Primary age modification | SBP, per +10 years | 0.05 (0.01 to 0.08) | Holm P=.009 | Change in P95-P50 ASD contrast |
| Primary age modification | SBP, joint spline × age | χ²=11.70, df=4 | .020 | n=153,359 |
| Primary age modification | DBP, age 35 | 0.00 (-0.07 to 0.07) | .995 | P95-P50 ASD contrast |
| Primary age modification | DBP, age 55 | 0.04 (-0.02 to 0.10) | .155 | P95-P50 ASD contrast |
| Primary age modification | DBP, age 75 | 0.08 (0.02 to 0.15) | .014 | P95-P50 ASD contrast |
| Primary age modification | DBP, per +10 years | 0.02 (-0.00 to 0.04) | Holm P=.051 | Change in P95-P50 ASD contrast |
| Primary age modification | DBP, joint spline × age | χ²=6.02, df=4 | .197 | n=153,286 |
| Post-index SMV modification | SBP | 0.10 (-0.12 to 0.32) | Holm P=.780 | Difference: SMV P90 minus P10 |
| Post-index SMV modification | SBP, joint spline × SMV | χ²=8.61, df=4 | .072 | n=67,438 |
| Post-index SMV ≤90-day interval sensitivity | SBP | 0.12 (-0.13 to 0.37) | .340 | Difference: SMV P90 minus P10 |
| Post-index SMV modification | DBP | -0.01 (-0.14 to 0.13) | Holm P=.906 | Difference: SMV P90 minus P10 |
| Post-index SMV modification | DBP, joint spline × SMV | χ²=5.27, df=4 | .260 | n=67,463 |
| Post-index SMV ≤90-day interval sensitivity | DBP | -0.06 (-0.22 to 0.09) | .409 | Difference: SMV P90 minus P10 |

Note. ASD denotes absolute successive difference, the interval-level contribution to average real variability. The primary BPV analysis allows intervals up to 365 days and adjusts for log1p days between outcome-specific measurements. Participant-level standard deviation (SD), coefficient of variation (CV) and average real variability (ARV) are descriptive. Intervals for the post-index SMV analysis were constructed within the post-index panel.

**Supplementary Figure S1. Temperature and blood-pressure curves across long-term climate context.**

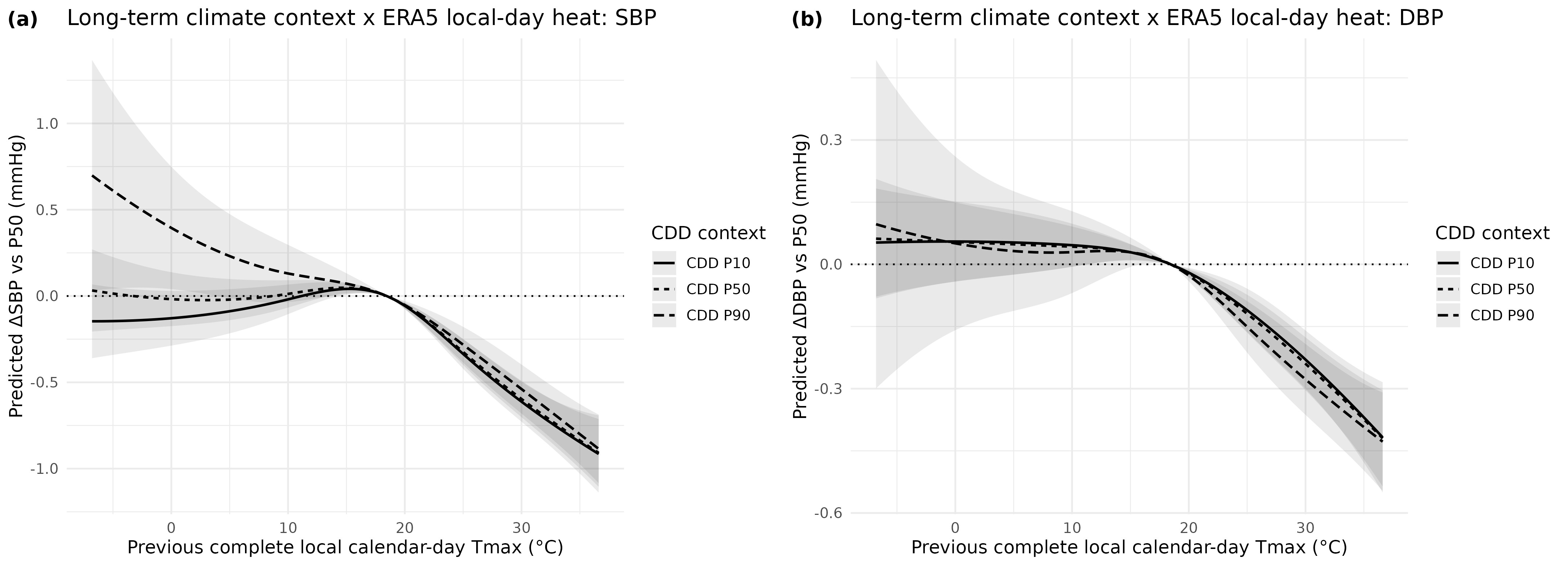

Note. Panels show (a) systolic and (b) diastolic model-implied differences relative to P50 across the P10, P50 and P90 of participant-level cooling-degree-day normal. Shaded bands show 95% confidence intervals.

**Supplementary Figure S2. Previous-local-day heat and visit-to-visit blood-pressure instability.**

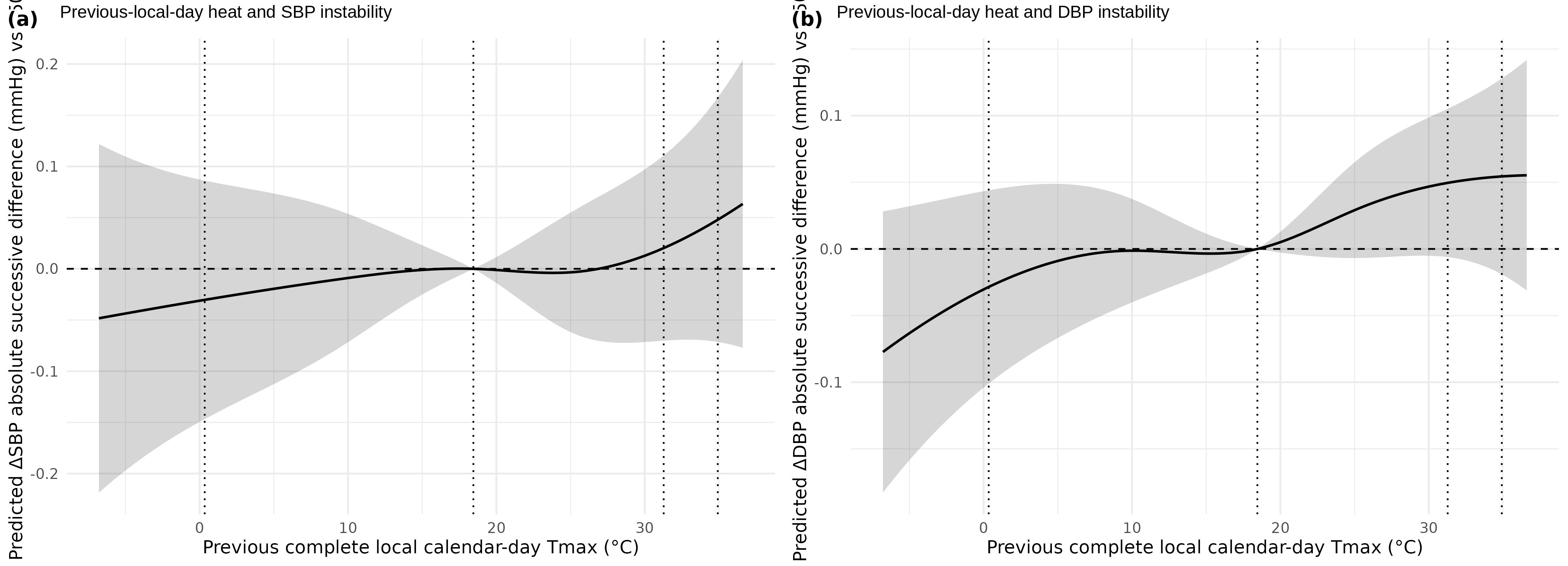

Note. Panels show model-implied changes in absolute successive difference for (a) systolic and (b) diastolic blood pressure relative to P50 previous-local-day Tmax. Shaded bands show 95% confidence intervals.
